# Genomic surveillance of antimicrobial resistant bacterial colonisation and infection in intensive care patients

**DOI:** 10.1101/2020.11.03.20224881

**Authors:** Kelly L. Wyres, Jane Hawkey, Mirianne Mirčeta, Louise M. Judd, Ryan R. Wick, Claire L. Gorrie, Nigel F. Pratt, Jill S. Garlick, Kerrie M. Watson, David V. Pilcher, Steve A. McGloughlin, Iain J. Abbott, Nenad Macesic, Denis W. Spelman, Adam W. J. Jenney, Kathryn E. Holt

## Abstract

**Background:** Third-generation cephalosporin-resistant Gram-negatives (3GCR-GN) and vancomycin-resistant Enterococci (VRE) are common causes of multi-drug resistant healthcare-associated infections, for which gut colonisation is considered a prerequisite. However, there remains a key knowledge gap about colonisation and infection dynamics in high-risk settings such as the intensive care unit (ICU), thus hampering infection prevention efforts.

**Methods:** We performed a three-month prospective genomic survey of infection and gut-colonisation with 3GCR-GN and VRE among patients admitted to an Australian ICU.

**Results:** Among 287 patients screened on admission, 17.4% and 8.4% were colonised by 3GCR-GN and VRE, respectively. *Escherichia coli* was the most common species (n=36 episodes, 58.1%) and the most common cause of 3GCR-GN infection. Only two VRE infections were identified. The rate of infection among patients colonised with *E. coli* was low, but higher than those who were not colonised on admission (6% vs 2% *E. coli, p*=0.3). While few patients were colonised with 3GCR-*Klebsiella pneumoniae* or *Pseudomonas aeruginosa* on admission (n=4), all such patients developed infections with the colonising strain. Genomic analyses revealed 10 putative nosocomial transmission clusters: four VRE, six 3GCR-GN, with epidemiologically linked clusters accounting for 21% and 6% of episodes, respectively (OR 4.3, *p*=0.02).

**Conclusions:** 3GCR-*E. coli* and VRE were the most common gut colonisers. *E. coli* was the most common cause of 3GCR-GN infection, but other 3GCR-GN species showed greater risk for infection in colonised patients. Larger studies are warranted to elucidate the relative risks of different colonisers and guide the use of screening in ICU infection control.

**Key points:** Surveillance for VRE or ESBL Gram-negative bacteria in ICU showed distinct species-specific clinical risk profiles. VRE and ESBL-*Escherichia coli* were the most common colonisers, but were associated with lower attack rate and transmission rate, compared with rarer coloniser *Klebsiella pneumoniae*.

## Introduction

Healthcare associated infections (HAI) result in considerable morbidity and mortality, with the total burden in high income countries exceeding that of influenza, tuberculosis and other major communicable diseases combined [1]. The emergence and spread of antimicrobial resistant (AMR) organisms, including difficult-to-treat multi-drug resistant (MDR) strains, has further exacerbated this problem [2], and as a consequence several MDR healthcare-associated bacterial pathogens are now recognised by the World Health Organization (WHO) as urgent threats to public health [3]. Among the WHO’s top priorities are carbapenem-resistant Gram-negatives (GNs, specifically *Enterobacteriaceae, Acinetobacter baumannii* and *Pseudomonas aeruginosa*) as well as vancomycin-resistant *Enterococci* (VRE).

In Australia, it is estimated that one in ten hospitalised patients suffers an HAI, and the most common MDR organisms are VRE, methicillin-resistant *Staphylococcus aureus* and extended-spectrum beta-lactamase (ESBL) producing *Enterobacteriaceae* [4]. Fortunately carbapenem-resistance has so far remained rare, although the prevalence has been slowly increasing [5]. Notably, Australia suffers a particularly high rate of vancomycin resistance among enterococcal infections (∼45% of *Enterococcus faecium* bacteremias [5]), primarily due to the endemic spread of healthcare-associated *E. faecium* sequence types (STs) 17, ST80 and ST796 [5–8].

Asymptomatic gut colonisation is considered a key risk factor for enterococcal, *Enterobacteriaceae* and other GN infections [9–12]. A number of hospitals have implemented rectal screening programs to identify patients colonised with VRE and/or carbapenemase-producing *Enterobacteriaceae* in high-risk wards such as intensive care units (ICUs), oncology and hematology wards [7,8,13,14]. However, few facilities conduct routine screening for ESBL-*Enterobacteriaceae* or other high-risk carbapenem-susceptible GNs in part due to a lack of evidence to inform the interpretation of these results and to appropriately target infection control measures. For example, only a small number of studies have investigated the risks associated with ESBL-*Enterobacteriaceae* colonisation at a species-specific level [9,11,12,15–18], but there is emerging evidence to suggest important variation, both in terms of the risk of infection and the risk of transmission in the hospital setting. We propose that a better understanding of these risks, as well as the broader colonisation and infection dynamics of AMR organisms in these settings, will facilitate better targeting of infection prevention and control practices.

Here we report a genome-resolved snapshot of high-risk AMR bacteria from rectal swabs and clinical specimens collected in an Australian ICU over a three-month period. We focus on GN organisms resistant to third-generation cephalosporins (including ESBL-*Enterobacteriaceae*) in addition to VRE, and leverage the genome data to perform high-resolution analysis of these bacterial populations.

## Methods

### Recruitment and specimen collection

The Alfred Hospital, Melbourne, Australia is a 350-bed tertiary referral hospital including a 45-bed ICU providing care for general medical and surgical patients plus specialist cardiac and trauma services. We conducted a prospective surveillance study for rectal colonisation of third generation cephalosporin-resistant GN (3GCR-GN) organisms and VRE in patients admitted to the ICU from Dec 21 2013 to 7 April 2014. This study was approved by the Alfred Health Human Research Ethics Committee, Project numbers #550/12 (19 February 2013) and #526/13 (10 December 2013). Patients aged ≥18 years and considered by nursing staff as likely to remain in the ICU >2 days were eligible for inclusion. The requirement for informed consent to participate was waived by the ethics committee as screening for AMR organisms is considered an infection control and surveillance activity, however patients were given the option to refuse screening. Rectal swabs were collected at time of recruitment (day 0-2 of ICU admission) and subsequently each 5-7 days during ICU stay. Information on age, sex, dates of hospital and ICU admission/s, surgery in the last 30 days, and antibiotic treatment in the last seven days were extracted from hospital records when swabs were taken. Dates of discharge and/or death were extracted from hospital records at the conclusion of the study. All clinical isolates recovered from ICU patients and identified as 3GCR-GN or VRE by the diagnostic laboratory as part of routine care were also stored for inclusion.

### Bacterial culture and sequencing

Gut colonising 3GCR-GN and VRE were identified by culture on ceftazidime and chromID VRE agar plates, respectively. Presumptive 3GCR-GN and VRE colonies were subjected to species identification via matrix-assisted laser desorption ionization time-of-flight analysis with a Vitek MS (bioMérieux). Clinical isolates were collected and identified via standard diagnostic protocols [10]. Genomic DNA was extracted and sequenced on the Illumina platform. Twenty-three isolates were selected for sequencing on the Oxford Nanopore MinION platform as described previously [19]. See **Supplementary Methods** for full details.

### Genomic analyses

Genomes were assembled using Unicycler v0.4.7 [20] and subjected to quality control (**Supplementary Methods**). Species were identified using Kraken v1.0 [21] and multi-locus sequence types (STs) were identified from assemblies using mlst (github.com/tseemann/mlst) where schemes were available (accessed from pubmlst.org [22]).

AMR genes were identified from final assemblies using Kleborate (github/katholt/kleborate), and these data used to calculate the predicted number of acquired AMR classes per isolate (defined as described in [23], **Supplementary Methods**). MDR was defined as acquired resistance for ≥3 drug classes [23].

Single nucleotide variants (SNVs) were identified by mapping Illumina reads to the relevant completed chromosomal reference genomes (ST-specific, **Table S1**) using Bowtie2 v2.2.9 [24] and SAMtools v1.9 [25], as implemented in the RedDog pipeline (github.com/katholt/RedDog) [26].

Illumina read data were deposited in the European Nucleotide Archive under project accessions PRJEB6891 and PRJNA646837, and hybrid genome assemblies were deposited in GenBank; accessions are listed in **Table S1** with isolate characteristics.

## Results

During the study period there were 770 ICU admissions, 31 patients with one or more 3GCR-GN infections (n=41 isolates), and two with VRE infections (n=2 isolates, both *E. faecium*, **Figure S1**). Of 430 patients eligible for participation in rectal screening, 66 declined to participate and 311 contributed one or more rectal swabs (72.3% of eligible non-refusers). The majority of participants were swabbed within the first two days of ICU admission (baseline screening swabs, n=287, 92.3%), including 79 with ≥1 additional follow-up swab (**Figure S1**). Participants with baseline swabs were 64.5% male (n=185), aged 18-93 years (median 57 years; IQR, 44-71 years, **Table S2**), and the majority (n=218, 76.0%) were known to have had recent healthcare exposure prior to ICU admission (surgical procedure within the last 30 days, transferred from another ward with first admission >2 days prior, or transferred from another hospital).

Third generation cephalosporin-resistant (3GCR) GN organisms were isolated from baseline screening swabs of 50 patients (17.4%, n=56 isolates), while VRE were isolated from 24 (8.4%, n=24 *E. faecium* isolates; **Figure S1**). Co-colonisation with 3GCR-GN and VRE was identified in 12 patients (4.2%), indicating a significant association between these organisms (OR 5.6, 95% CI 2.2-15.5, p=0.0001 using Fisher’s Exact Test). There were no significant differences in colonisation rates between males and females, and no association between age and colonisation status, with the exception of VRE in females (median age 73.5 years amongst carriers versus 55.5 years amongst non-carriers, *p*=0.04 using Wilcoxon Rank Sum test; **Figure 1**). Neither surgery within the last 30 days, recent healthcare exposure (defined as above), or antibiotic treatment within the last seven days, were significantly associated with 3GCR-GN or VRE colonisation at baseline (using univariate and multivariate logistic regression, with age and sex as covariates; **Table S3**).

**Figure 1:**
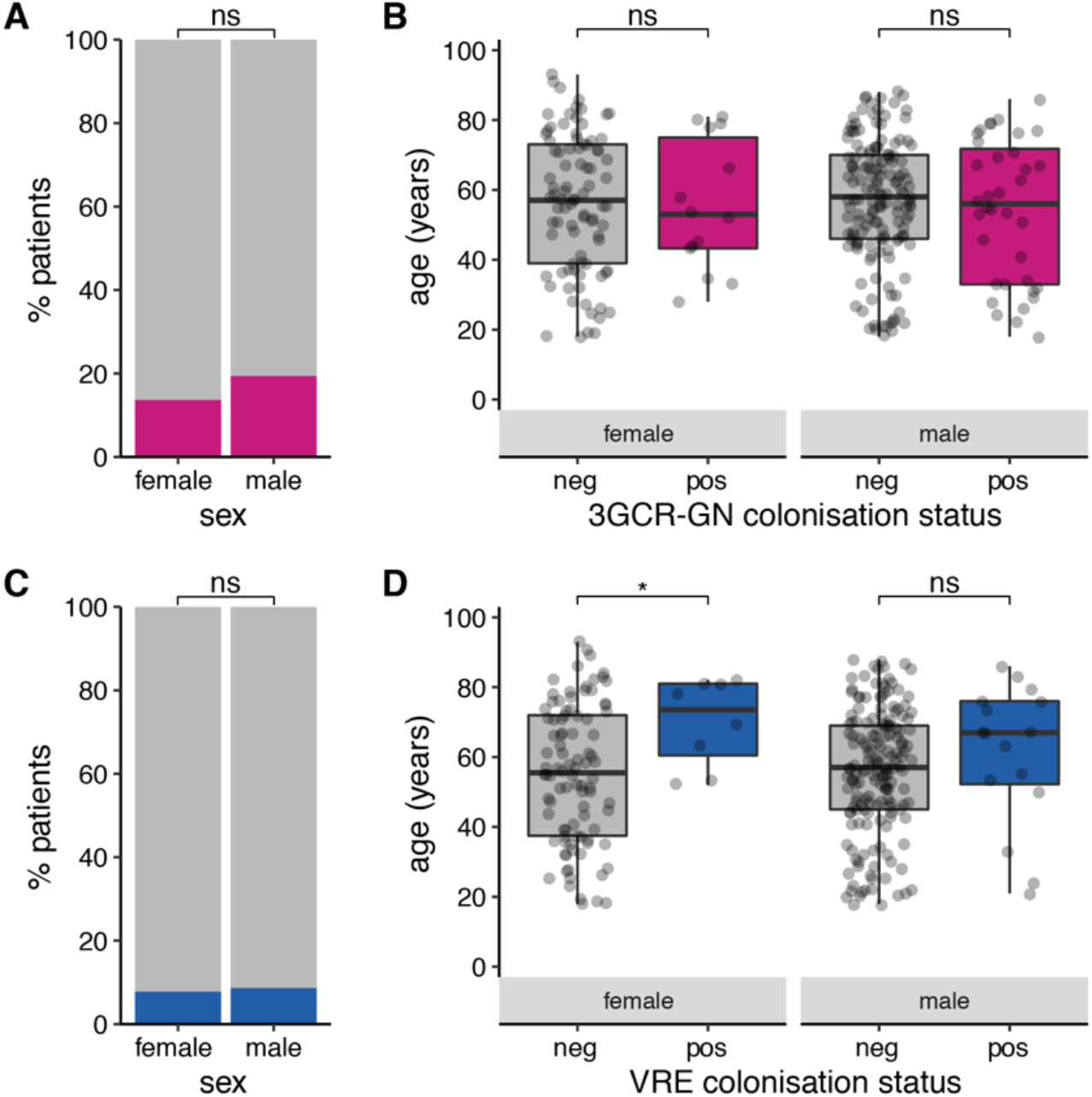
Prevalence of 3GCR-GN and VRE gut colonisation on admission to the ICU. Prevalence of third generation cephalosporin-resistant Gram-negative (3GCR-GN) organisms (**A**) and VRE organisms (**C**) among males and females for whom rectal swabs were collected within two days of ICU admission. Age distributions are shown for culture-confirmed 3GCR-GN (**B**) and VRE (**D**) carriers and non-carriers, stratified by sex. ns, non-significant using Fisher’s Exact Test (**A, C**) or Wilcoxon Rank Sum test (**B, D**); *, p< 0.05.

A total of 138 rectal swabs were collected from 103 patients between three days and 12 weeks after ICU admission (**Table S4, Figure S2**). Twenty-five swabs (18.1%) from 18 patients (17.5%) were positive for ≥1 3GCR-GN organism (n=30 isolates); 15 swabs (10.9%) from ten patients (9.7%) were positive for VRE (n=15 isolates). Notably, 3GCR-GN organisms were cultured from 15/61 patients (24.6%) who were culture-negative for 3GCR-GN colonisation at baseline, and VRE were cultured from 13/75 (17.3%) patients who were culture-negative for VRE colonisation at baseline, consistent with acquisition and/or overgrowth of the organisms during ICU stay.

Among the 127 3GCR-GN and 41 VRE isolates cultured from ICU patients, 112 (88.2%) and 41 (100%) respectively were successfully revived and sequenced (**Supplementary Methods, Figure S1**). We combined clinical and genome data to identify distinct colonisation and infection episodes, defined as unique combinations of species and ST (derived from whole-genome sequences (WGS)), per patient and specimen type. This identified 33 3GCR-GN and two VRE infection episodes (**Table 1**). The latter included one urinary tract infection and one respiratory infection. 3GCR-GN infections were mainly respiratory (n=19, 57.6%), wound (n=5, 15.2%) and bloodstream (n=4, 12.1%) infections, and the most common agents were *Escherichia coli* (10 episodes, 30.3%) and *Klebsiella pneumoniae* (6 episodes, 18.2%; **Table 1, Figure 2**). *E. coli* was also the most common 3GCR-GN gut coloniser (36/62 3GCR-GN colonisation episodes, 58%), followed by *Enterobacter hormaechei, K. pneumoniae, Klebsiella aerogenes*, and *Klebsiella oxytoca* (four episodes, 6.5%, each, **Table 1**). Thirty-three unique VRE colonisation episodes were identified. Overall, 88 patients were colonised and/or infected with 3GCR-GN and/or VRE. Most (62, 70.5%) experienced a single episode of either 3GCR-GN (n=61) or VRE (n=1), however 17 patients (20.2%) had two episodes (16 patients with two different species/STs, 13 with one VRE and one 3GCR-GN species) and nine patients (10.1%) had ≥3 episodes (seven with ≥3 different species/STs, including five with VRE).

**Table 1:** Colonisation and infection episodes by species.

| Species | Total colonisation episodes (n baseline) | Infection episodes |  |  |  | *p*-value |
| --- | --- | --- | --- | --- | --- | --- |
|  |  | Total | Pt colonised with same strain (n baseline) | Pt colonised at baseline (%) | Pt not colonised at baseline (%) |  |
| **VRE** |  |  |  |  |  |  |
| *Enterococcus faecium* | 33 (25) | 2 | 0 | 0 | 1 (0.4%) | - |
| **3GC-resistant GN** |  |  |  |  |  |  |
| *Escherichia coli* | 36 (33) | 10 | 1 (0) | 2 (6%) | 4 (2%) | 0.2926 |
| *Klebsiella pneumoniae* | 4 (2) | 7 | 3 (2) | 2 (100%) | 3 (1%) | $1.72 \times 10^{-15}^{**}$ |
| *Pseudomonas aeruginosa* | 3 (2) | 2 | 1 (1) | 2 (100%) | 0 | $< 2.2 \times 10^{-16}^{**}$ |
| *Enterobacter hormaechei* | 4 (3) | 3 | 0 | 0 | 1 | - |
| *Klebsiella aerogenes* | 4 (1) | 3 | 0 | 0 | 1 | - |
| *Enterobacter asburiae* | 0 | 2 | 0 | 0 | 1 | - |
| *Stenotrophomonas maltophilia* | 0 | 2 | 0 | 0 | 1 | - |
| *Serratia marcescens* | 1 (0) | 2 | 0 | 0 | 0 | - |
| *Achromobacter xylosoxidans* | 0 | 1 | 0 | 0 | 1 | - |
| *Burkholderia cenocepacia* | 0 | 1 | 0 | 0 | 1 | - |
| *Citrobacter freundii* | 2 (2) | 1 | 0 | 0 | 0 | - |
| *Citrobacter portucalensis* | 1 (1) | 0 | 0 | 0 | 0 | - |
| *Enterobacter kobei* | 1 (0) | 0 | 0 | 0 | 0 | - |
| *Escherichia marmotae* | 1 (1) | 0 | 0 | 0 | 0 | - |
| *Klebsiella oxytoca* | 4 (4) | 0 | 0 | 0 | 0 | - |
| *Morganella morganii* | 1 (0) | 0 | 0 | 0 | 0 | - |

**\*\*** $p < 0.00005$ for association between colonisation at baseline and infection with same species (proportion test).
Distinct episodes of colonisation and infection were defined as those with a unique combination of patient ID, specimen type, species and multi-locus sequence type. Colonisation episodes are expressed as total detected and number detected at baseline (within first two days after ICU admission). Strains were defined on the basis of chromosomal SNVs ( $\leq 7$ SNVs between isolates). Pt, patient; VRE, vancomycin-resistant Enterococci.

**Figure 2:**
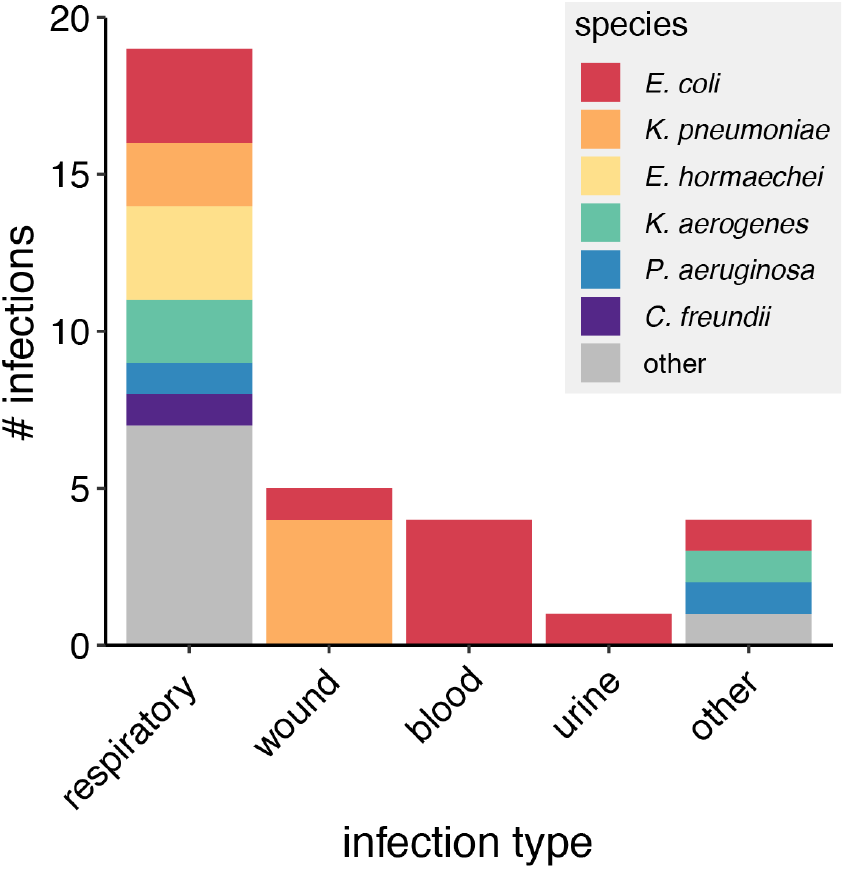
Infections caused by 3GCR-GN organisms. Bar chart shows the number of distinct respiratory, wound, blood, urine and other infections by species as indicated in the legend. Distinct infections were defined as those represented by unique combinations of species, multi-locus sequence type, patient and specimen type.

The majority of 3GCR-GN colonisation and infection isolates were predicted to be MDR (see **Methods**): n=44/61 (72%) distinct colonisation episodes and n=18/29 (62%) distinct infection episodes. MDR was particularly common among *E. coli* (n=44 episodes, 96%), *K. pneumoniae* (n=9, 90%) and *Serratia marcescens* (n=3, 100%). WGS identified acquired ESBL genes in 20 representative 3GCR-GN colonisation (32.3%) and 12 representative 3GCR-GN infection (36.4%) isolates. These were mostly MDR *K. pneumoniae* carrying *bla*_CTX-M-15_ (n=8) or MDR *E. coli* carrying diverse *bla*_CTX-M_ genes (n=19, including *bla*_CTX-M-14_, *bla*_CTX-M-15_, *bla*_CTX-M-24-like_, *bla*_CTX-M-27_, *bla*_CTX-M-62_). Carbapenemase genes were rare: two patients had wound or tissue infections with *K. pneumoniae* ST231 carrying *bla*_OXA-48,_ one of whom was also colonised by *K. pneumoniae* ST231 with *bla*_OXA-48._ A third patient was colonised with *S. marcescens* carrying *bla*_IMP-4_.

Most VRE isolates representing distinct episodes (n=32, 91%) carried the *vanB* vancomycin resistance operon and the remainder (n=3, 9%) the *vanA* operon, which confers resistance to vancomycin plus teicoplanin [27]. Additionally, 32 of these isolates (91%) were predicted to be MDR due to the presence of genes conferring resistance to tetracyclines and/or high-level resistance to gentamicin, streptomycin and/or streptogramins.

We assessed species-specific infection rates amongst patients testing culture-positive at baseline screening, versus those testing culture-negative, for all species with ≥2 infections identified amongst patients with baseline screening data (*E. coli, K. pneumoniae, P. aeruginosa*). Infection rates were higher amongst baseline-culture-positive patients, although the difference was statistically significant only for *K. pneumoniae* and *P. aeruginosa* (**Table 1**). There were few patients for whom WGS data were available for isolates of the same species detected in baseline screening and infections (two patients for each species). For n=2/2 such cases with *K. pneumoniae* (ST231, ST323) and n=1/2 with *P. aeruginosa* (ST357), the WGS data confirmed the infections were caused by the colonising strain (0-7 pairwise SNVs), whereas both *E. coli* infections were caused by different strains to those detected on baseline screening swabs (different STs; **Table 1, Supplementary Results**). A further two patients tested culture-negative on baseline screening, but subsequently tested positive on follow-up screening for the same strain that was isolated from their diagnostic specimens (*K. pneumoniae* ST323, *E. coli* ST393; **Supplementary Results**).

Finally, we assessed evidence for nosocomial transmission using WGS and patient data. The isolates were diverse, with 62 STs identified amongst 3GCR-GN and 8 amongst VRE. Most 3GCR-GN STs (n=48, 77%) were unique to a single patient; however, ten *E. coli* STs (42%), two *K. pneumoniae* (40%), one *K. oyxtoca* (33%), one *E. hormachaei* (17%) and five VRE STs (63%) were found in multiple patients (**Figure 3a**). We defined probable strain transmission events on the basis of chromosomal SNVs (see **Methods, Figure 3b-c**). Using a threshold of ≤20 SNVs for transmission of 3GCR-GN (based on recent studies [10,13,14,28]) we identified six putative 3GCR-GN transmission clusters each involving 2-3 patients (maximum 8 SNVs, **Table 2**). With a single exception, all patients within clusters were epidemiologically linked (overlapping ICU stays or ≤14 days between stays). Assuming one patient in each epidemiologically linked cluster was the index patient, the data suggest only six (6.3%) 3GCR-GN episodes resulted from recent nosocomial transmission (n=2, 6.1% of infections; n=4, 6.5% of colonisation). We also identified *K. pneumoniae* ST323 isolates from two patients that differed by 45-47 SNVs (**Figure 3a**), suggesting recent acquisition from a common unsampled source and consistent with previous data showing that *K. pneumoniae* ST323 were circulating more broadly within the hospital during the study period [29].

**Table 2:**
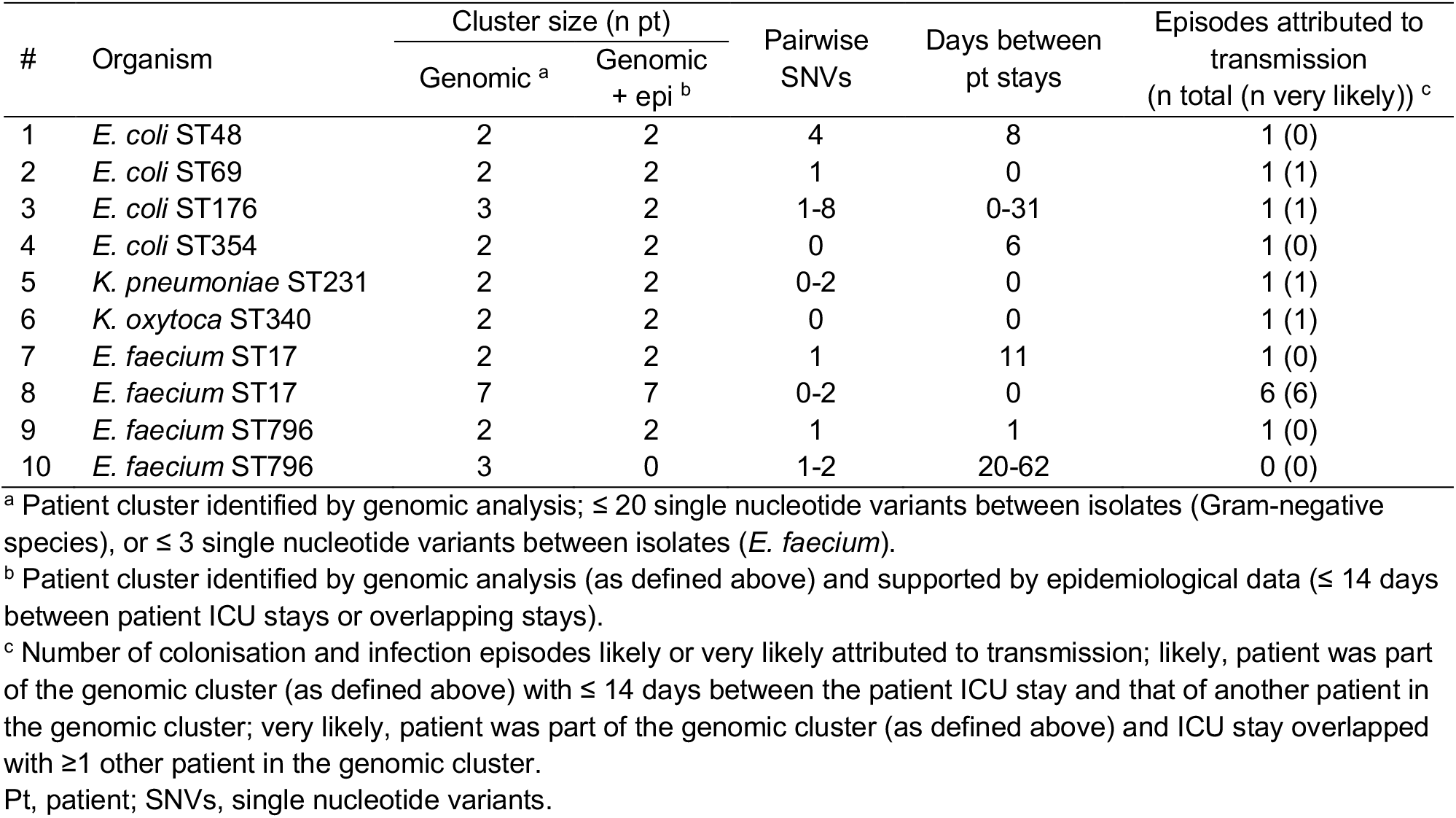
Putative nosocomial transmission clusters.

| # | Organism | Cluster size (n pt) |  | Pairwise SNVs | Days between pt stays | Episodes attributed to transmission (n total (n very likely))<sup>c</sup> |
| --- | --- | --- | --- | --- | --- | --- |
|  |  | Genomic<sup>a</sup> | Genomic + epi<sup>b</sup> |  |  |  |
| 1 | *E. coli* ST48 | 2 | 2 | 4 | 8 | 1 (0) |
| 2 | *E. coli* ST69 | 2 | 2 | 1 | 0 | 1 (1) |
| 3 | *E. coli* ST176 | 3 | 2 | 1-8 | 0-31 | 1 (1) |
| 4 | *E. coli* ST354 | 2 | 2 | 0 | 6 | 1 (0) |
| 5 | *K. pneumoniae* ST231 | 2 | 2 | 0-2 | 0 | 1 (1) |
| 6 | *K. oxytoca* ST340 | 2 | 2 | 0 | 0 | 1 (1) |
| 7 | *E. faecium* ST17 | 2 | 2 | 1 | 11 | 1 (0) |
| 8 | *E. faecium* ST17 | 7 | 7 | 0-2 | 0 | 6 (6) |
| 9 | *E. faecium* ST796 | 2 | 2 | 1 | 1 | 1 (0) |
| 10 | *E. faecium* ST796 | 3 | 0 | 1-2 | 20-62 | 0 (0) |

<sup>a</sup> Patient cluster identified by genomic analysis; $\leq 20$ single nucleotide variants between isolates (Gram-negative species), or $\leq 3$ single nucleotide variants between isolates (*E. faecium*).
<sup>b</sup> Patient cluster identified by genomic analysis (as defined above) and supported by epidemiological data ( $\leq 14$ days between patient ICU stays or overlapping stays).
<sup>c</sup> Number of colonisation and infection episodes likely or very likely attributed to transmission; likely, patient was part of the genomic cluster (as defined above) with $\leq 14$ days between the patient ICU stay and that of another patient in the genomic cluster; very likely, patient was part of the genomic cluster (as defined above) and ICU stay overlapped with $\geq 1$ other patient in the genomic cluster.
Pt, patient; SNVs, single nucleotide variants.

**Figure 3:**
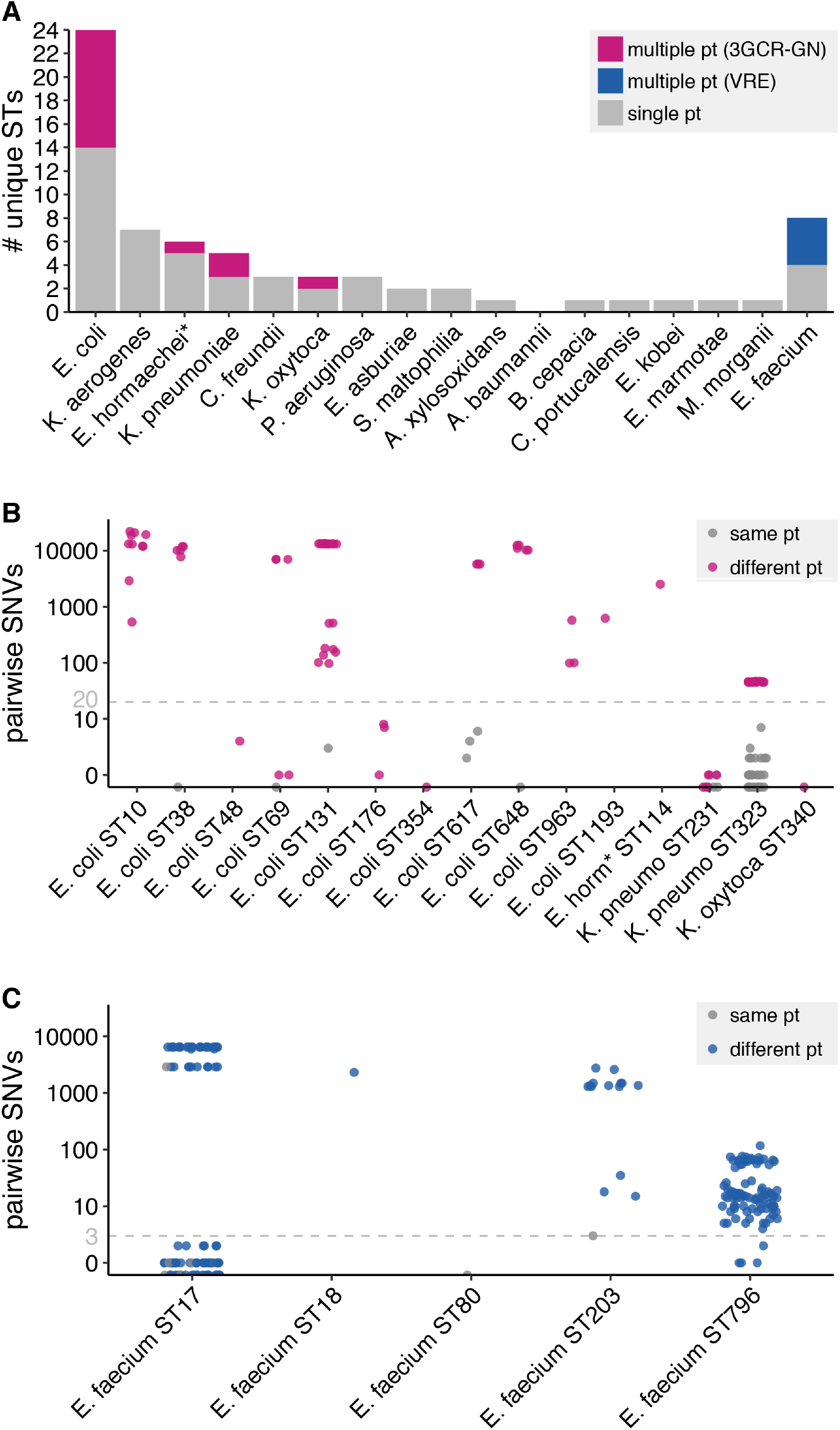
3GCR-GN and VRE strain diversity. (**A**) Bar plot showing the number of unique multi-locus sequence types (STs) identified for each third-generation cephalosporin-resistant Gram-negative (3GCR-GN) species plus vancomycin-resistant *E. faecium* (VRE) Bars are coloured as indicated in the legend. **(B, C)** Pairwise single nucleotide variants (SNVs) for pairs of isolates assigned to the same ST for 3GCR-GN species (**B**) and VRE (**C**). Grey points indicate pairs of isolates from the same patient (pt), pink and blue points indicate pairs of isolates from different patients for 3GCR-GN and VRE isolates, respectively. Grey dashed lines indicate the SNV thresholds used to define putative transmissions (n≤20 SNVs for 3GCR-GN and n≤3 SNVs for VRE, see **Results** for details). \**E. hormaechei* multi-locus sequence typing was performed using the *Enterobacter cloacae* scheme, which covers both species.

Interpretation of VRE pairwise SNVs is more complex because several healthcare-associated lineages are known to be circulating in Melbourne and the regional hospital network. Consequently, it is not uncommon to find near-identical isolates in patients from different wards, hospitals or cities without epidemiological links [7,8,13]. We therefore used a more conservative threshold of ≤3 SNVs to define putative recent VRE transmissions (based on empirical distribution, see **Figure S3**), which identified four clusters of 2-7 patients each (**Table 2**). Two clusters were ST17 (both epidemiologically linked clusters, separated by >2800 SNVs) and two were the locally emerged ST796 [30,31] (one epidemiologically linked cluster, separated by 4-6 SNVs). Assuming one patient in each epidemiologically linked cluster is the index patient, we estimate eight (28.6%) VRE episodes resulted from recent transmission (n=1 infection and n=7, 21.2% of colonisation episodes). This should be considered a conservative estimate considering the difficulty in distinguishing clusters, particularly for ST796. Nevertheless, the transmission burden is significantly greater than that estimated for 3GCR-GN organisms (OR=4.3, 95% CI 1.2-16.6, *p*=0.02 using Fisher’s Exact test).

## Discussion

We have presented a holistic investigation of 3GCR-GN and VRE colonising and infecting patients in an Australian ICU over a three-month duration. Approximately 17% patients were colonised by at least one 3GCR-GN at ICU admission. Just under 40% of these isolates were detected as ESBL-*Enterobacteriaceae*, hence our data are comparable to reports from Europe where 7-13% of ICU-admitted patients were colonised with these organisms [15,17,32]. Additionally, 9% of patients were colonised with VRE at ICU admission; a similar prevalence to that reported recently for patients in a hospital network in Singapore [33], but lower than the 17.5% (95% CI 13.7-21.9) indicated in a previous point-prevalence survey of patients in our hospital [34].

*E. coli* was the dominant colonising and infecting organism, and has been previously reported as the most common cause of GN blood-stream infections in Australia [5]. These findings are consistent with the notion of colonisation as a prerequisite for disease and the logic of scales, whereby the most common colonisers have more opportunities to cause infection and therefore contribute the greatest burden of disease. Our data indicated a higher prevalence of infections among patients colonised at admission with 3GCR *E. coli, K. pneumoniae* or *P. aeruginosa* than those who were not colonised. However, there were stark differences in the rate of infection; just 6% for *E. coli*, versus 100% (n=2/2 each) for *K. pneumoniae* and *P. aeruginosa*. While these data must be interpreted with caution due to the small sample size, we note that the trends are consistent with previous reports [9,15] including a large study of 2386 patients colonised by ESBL-*K. pneumoniae* or ESBL-*E. coli*, which found that the former were twice as likely as the latter to develop an infection with the same organism (6.8% versus 3.2% [9]).

The inclusion of WGS data also allowed us to investigate putative transmissions, identifying three VRE and six 3GCR-GN epidemiologically linked transmission clusters. The estimated burden of all VRE and 3GCR-GN episodes attributable to transmission was 21.2% and 6.3%, respectively (OR 4.3, p=0.0209). The VRE clusters involved ST17 and ST796, which are considered healthcare-adapted lineages and are widely distributed in Australia [5–8,31,35].

While four of the six 3GCR-GN transmission clusters involved *E. coli*, these included just four of 24 distinct *E. coli* STs, and did not include the globally distributed ST131, ST10 or ST38. This is consistent with a recent report showing that these *E. coli* STs were commonly identified from patients in other Melbourne hospitals, but rarely associated with nosocomial transmission [13]. Hence our data adds to the growing evidence base [13,16–18,36] that ESBL-*E. coli* are mainly spread in the community setting.

While the risk of ESBL-*E. coli* transmission in hospitals may be low, there is emerging evidence that the risk may be 2-4 times greater for other ESBL-*Enterobacteriaceae* such as *K. pneumoniae* and *Enterobacter* sp. [17,18]. In our study, 2/11 (18.2%) ESBL-*K. pneumoniae* episodes were attributed to recent transmission, with one additional episode attributed to broader transmission within the hospital as reported previously [29]; however, a larger sample size is required to make a definitive comparison.

The inclusion of contemporaneous rectal screening and clinical isolates is a key strength of this study, which in combination with the breadth of species sampling and use of WGS, has allowed high-resolution analysis of the dynamics of AMR organisms in the ICU. Notably, the majority of putative transmissions we identified would have been missed if not for the inclusion of rectal screening. However, the study also has several limitations: firstly, we were not able to screen all eligible patients, nor to follow our patients beyond the three-month study period, thus we likely underestimate the true burdens of infection and transmission. Secondly, due to its short duration this study was underpowered for the assessment of patient risk factors (e.g. recent surgery or antibiotic use), which we expect to impact the risk of infection and onwards transmission. Thirdly, our focus on the ICU as a high-risk setting for HAI fails to capture the additional burden of 3GCR-GN and VRE in other wards. Nevertheless, the results highlight and contrast several key features of 3GCR-GN and VRE dynamics. Importantly, the data suggest the most common AMR colonisers (VRE and *E. coli*) do not necessarily pose the greatest risk to patients. A more detailed assessment of species-specific risks (especially attack rate and risk of transmission) is needed to guide the efficient use of rectal screening programs and targeted approaches to infection prevention and control aimed at limiting the MDR disease burden.

## Data Availability

All data supporting the manuscript are provided within the main document and/or supplementary materials, or via public data repositories (European Nucleotide Archive, Genbank and/or Figshare) as specified within the manuscript materials.

https://doi.org/10.26180/c.5194736

## Acknowledgements

We gratefully acknowledge the contribution and support of Janine Roney, Mellissa Bryant, Jennifer Williams, and Noelene Browne at the Alfred Hospital, and the sequencing team at the Wellcome Trust Sanger Institute. We also acknowledge the curators of the relevant MLST schemes and the developers of the PubMLST website (https://pubmlst.org/ Jolley and Martin Maiden. The development of that website was funded by the Wellcome Trust.

## Supplementary Methods

### Bacterial culture and sequencing

Rectal swabs were plated onto ceftazidime agar plates to identify third-generation cephalosporin-resistant Gram-negative (3GCR-GN) organisms and chromID VRE agar to identify vancomycin-resistant Enterococci (VRE) (BioMérieux, Marcy L’Etoile, France). Additionally, ciprofloxacin agar plates were used to determine if the Gram-negative organisms were resistant to fluoroquinolones. Plates were incubated in air at 36°C for 24-48 hours. Following species identification, purified cultures were stocked in glycerol and stored at −80°C. Clinical isolates were collected and identified via standard diagnostic protocols.

Genomic DNA was extracted from overnight cultures using a phenol:chloroform protocol and phase lock gel tubes as previously described [1]. Paired-end sequencing libraries were prepared using the Nextera XT kit (Illumina Inc) and sequenced on either the Illumina HiSeq platform (3GCR-GN isolates, 125 bp paired-end reads) or the Illumina MiSeq platform (VRE isolates, 300 bp paired-end reads). Isolates representing multi-locus sequence types of interest (those identified from ≥2 patients, or for which gut colonising and infecting isolates were identified from the same patient) were selected for sequencing on the Oxford Nanopore MinION R9.4.1 flow cell (n = 23 isolates, one for each sequence type of interest, see **Table S1**) [2].

### Genome assembly and quality control

Draft genomes were assembled using SPAdes v3.12.0 [3] optimised in Unicycler v0.4.7 [4] (using default parameters, and separately using the “--depth_filter 0.0” to allow for assessment of contamination). Illumina data were subjected to quality control to exclude low quality genomes if any of the following thresholds were met: 1) estimated depth <20x; 2) total assembly length (from default parameters) >9 Mbp or 3.5 Mbp for 3GCR-GN and VRE, respectively; 3) total assembly length (from default parameters) >6.5 Mbp or 3 Mbp and total contig count >500 for 3GCR-GN and VRE, respectively; 4) total number of assembly graph dead-ends >10 or >110 for 3GCR-GN and VRE, respectively and combined length of contaminant contigs >0.5% genome length; 5) combined length of contaminant contigs >0.5% genome length and contaminants identified as the same species as the dominating organism using Kraken v1.0 [5] (in such a case the contaminants were considered likely to impact downstream mapping and variant-calling analyses, whereas contaminants originating from a different species were unlikely to map to the reference chromosome). Contaminant contigs were identified from the “--depth_filter 0.0” assembly (defined as mean contig depth <0.25x chromosomal depth) and these were filtered out of the default-parameter assemblies that were used for downstream analyses.

For 23 isolates subjected to additional long-read sequencing, completed genome assemblies were generated using Unicycler v0.4.7 [4]. Two VRE assemblies failed to resolve the chromosome into circular replicons via this method, and were assembled independently using Flye v2.6 [6] to perform long-read-only assemblies that were polished with the Illumina reads using Pilon v1.22 [7]. Genome assemblies have been deposited in Genbank (accession numbers listed in **Table S1**) and are available via Figshare at https://doi.org/10.26180/c.5194736.

### Antimicrobial resistance profiles

Antimicrobial resistance (AMR) genes were identified from genome assemblies using Kleborate (github/katholt/kleborate) which employs a curated version of the CARD AMR gene database v3.0.8 [8]. These data were combined with phenotype data based on the ceftazidime and ciprofloxacin plate growth (3GCR-GN only) or chromID VRE plate growth (VRE only) to calculate the predicted number of acquired AMR classes per isolate, defined on a species-specific basis as described in [9] (acquired resistances could not be determined for six isolates for which species definitions were unavailable, see **Table S1**). High-level resistances for gentamicin, streptomycin and streptogramins were predicted for *E. faecium* genomes on the basis of the relevant acquired genes described in [10].

## Supplementary Results

### Comparison of colonising and infecting isolates from the same patient

We identified patients who had infection and colonisation episodes caused by the same species and sequence type (ST). Overall, four of 36 patients colonised by *E. coli* (11.1%) also had an *E. coli* infection but only one of these was caused by an isolate sharing the same ST as the colonising strain (2.7%). Similarly, one of 33 patients colonised by VRE had a VRE infection and this was caused by an isolate of the same ST (3.0%). In contrast, three of four patients colonised by *K. pneumoniae* (75.0%) and two of three patients colonised by *P. aeruginosa* (66.6%) also had infections caused by isolates sharing the same ST as the colonising strain.

The groups of matching colonisation and infection isolates comprised *E. coli* ST393, *K. pneumoniae* ST323 (two patients), *K. pneumoniae* ST231, *P. aeruginosa* ST357, *P. aeruginosa* ST471 and VRE ST17. We generated high quality completed reference genomes for one representative isolate of each of these STs (see **Table S1**). We then compared all available isolates from each of the case patients to their respective ST reference genomes, which confirmed that the *E. coli*, the *P. aeruginosa* ST357 and the three *K. pneumoniae* infections were caused by strains that were very closely related to those isolated from rectal carriage in the same patient (0, 0 and 0-7 chromosomal SNVs between pairs of infection and carriage isolates, respectively). These included the two *K. pneumoniae* infections (one ST231 and one ST323) and the *P. aeruginosa* infection (ST357) reported in **Results** among patients colonised by the same strain at baseline. The second patient harbouring *K. pneumoniae* ST323 was negative for rectal colonisation at base-line screening, but was positive from a rectal swab and a sputum sample collected 39 days later. The patient carrying *E. coli* ST393 was negative for rectal colonisation at baseline but positive from a subsequent rectal swab collected after a positive blood culture. The pair of VRE ST17 isolates from patient AH0390 (one from a rectal swab collected >2 days after admission and one causing empyema) differed by 2894 SNVs, suggesting that these were unrelated episodes. Supporting this hypothesis, our subsequent transmission analyses indicated that the infecting isolate was part of a transmission cluster and was not the index case, whereas the carriage isolate was not part of any transmission cluster. Comparison of the two *P. aeruginosa* ST471 isolates from patient AH0296 (one baseline rectal carriage and one respiratory infection isolate) suggested that these were independent (458 SNVs). Review of the patient record indicated that AH0296 was a cystic fibrosis patient with chronic *Pseudomonas* sp. colonisation; hence it is possible that the rectal carriage and sputum isolates represent divergent descendants from a single common ancestor that was acquired months or years prior to this study.

## Supplementary Tables

**Table S1: Specimen and genotype information of isolates included in this study.**

See separate Excel sheet, also available at https://doi.org/10.26180/c.5194736.

**Table S2:** Characteristics of the patient cohort subjected to rectal screening within the first two days of ICU admission.

|  | N Participants (%) |
| --- | --- |
| Sex (male) | 185 (64.5) |
| Age (years): |  |
| 18-29 | 32 (11.2) |
| 30-39 | 27 (9.4) |
| 40-49 | 36 (12.5) |
| 50-59 | 62 (21.6) |
| 60-69 | 49 (17.1) |
| 70-79 | 52 (18.1) |
| >80 | 29 (10.1) |
| Antibiotics last 7 days | 219 (76.3) |
| Surgery last 30 days | 177 (61.7) |
| Recent healthcare exposure<sup>a</sup> | 218 (76.0) |

<sup>a</sup> surgical procedure within the last 30 days, transferred from another ward in the Alfred hospital with first admission >2 days prior and/or transferred from another hospital.

**Table S3:** Logistic regression for factors associated with colonisation (culture-positive rectal swab) at baseline screening (day 0-2 of ICU admission)

|  | 3GCR-GN | VRE |
| --- | --- | --- |
| **Univariate analysis** |  |  |
| Sex (male) | 0.42 (p=0.22) | 0.11 (p=0.81) |
| Age | -0.01 (p=0.41) | 0.03 (p=0.04)* |
| Age (within males) | -0.01 (p=0.38) | 0.02 (p=0.31) |
| Age (within females) | -0.00 (p=0.85) | 0.04 (p=0.05) |
| Antibiotics last 7 days | 0.58 (p=0.16) | 0.48 (p=0.40) |
| Surgery last 30 days | -0.29 (p=0.37) | 0.24 (p=0.60) |
| Recent healthcare exposure<sup>a</sup> | -0.13 (p=0.72) | 0.85 (p=0.18) |
| **Multivariate analysis** |  |  |
| Sex (male) | 0.48 (p=0.17) | 0.14 (p=0.77) |
| Age (years) | -0.01 (p=0.54) | 0.03 (p=0.04)* |
| Antibiotics last 7 days | 0.77 (p=0.08) | 0.50 (p=0.41) |
| Surgery last 30 days | -0.51 (p=0.13) | -0.00 (p=1.00) |

<sup>a</sup> surgical procedure within the last 30 days, transferred from another ward in the Alfred hospital with first admission >2 days prior and/or transferred from another hospital.
3GCR-GN, third-generation cephalosporin-resistant Gram-negative; VRE, vancomycin resistant Enterococci; \*, $p \leq 0.05$ ; \*\* $p < 0.0005$ .

**Table S4:** Prevalence of third generation cephalosporin-resistant Gram-negatives (3GCR-GN) and vancomycin-resistant *E. faecium* (VRE) isolated from rectal screening swabs.

| Time since admission (days) | Swabs processed (n) | VRE positive (n) | VRE prevalence (%) | 3GCR-GN positive (n) | 3GCR-GN prevalence (%) | Total GN organisms (n) |
| --- | --- | --- | --- | --- | --- | --- |
| 0-2 | 287 | 24 | 8.4 | 50 | 17.4 | 63 |
| 3-7 | 84 | 6 | 7.1 | 8 | 9.5 | 10 |
| 7-14 | 28 | 2 | 7.1 | 7 | 25.0 | 8 |
| 15-21 | 12 | 3 | 25.0 | 2 | 16.7 | 1 |
| 22-78<sup>a</sup> | 14 | 4 | 28.6 | 8 | 57.1 | 11 |

## Supplementary Figures

**Figure S1:**
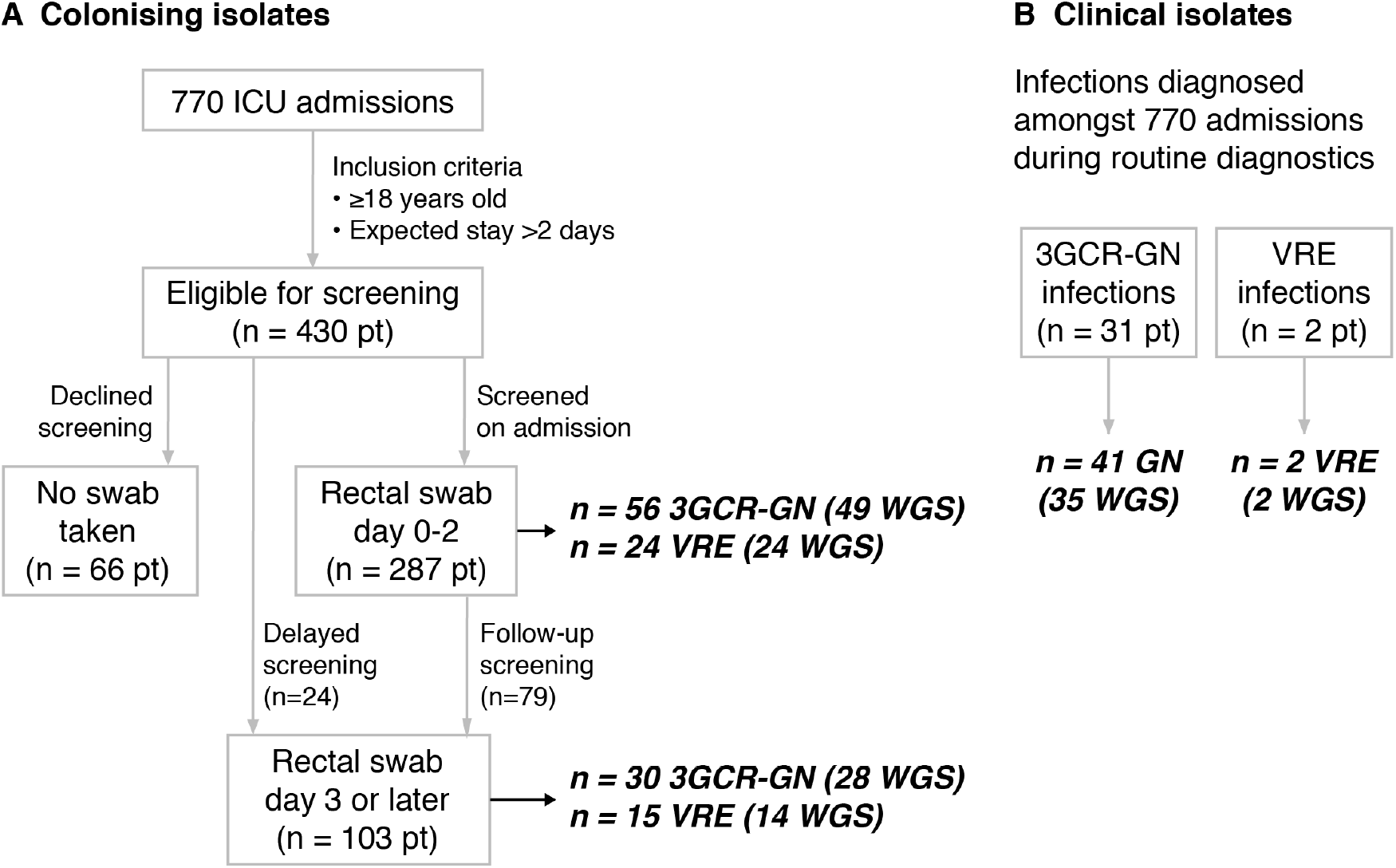
Sources of (A) colonising and (B) clinical isolates from ICU patients. 3GCR-GN, third generation cephalosporin-resistant Gram-negatives; VRE, vancomycin-resistant enterococci; pt patients; WGS, whole-genome sequences. Bold italics indicates number of isolates, numbers in parentheses indicate the number of WGS included in analyses.

**Figure S2:**
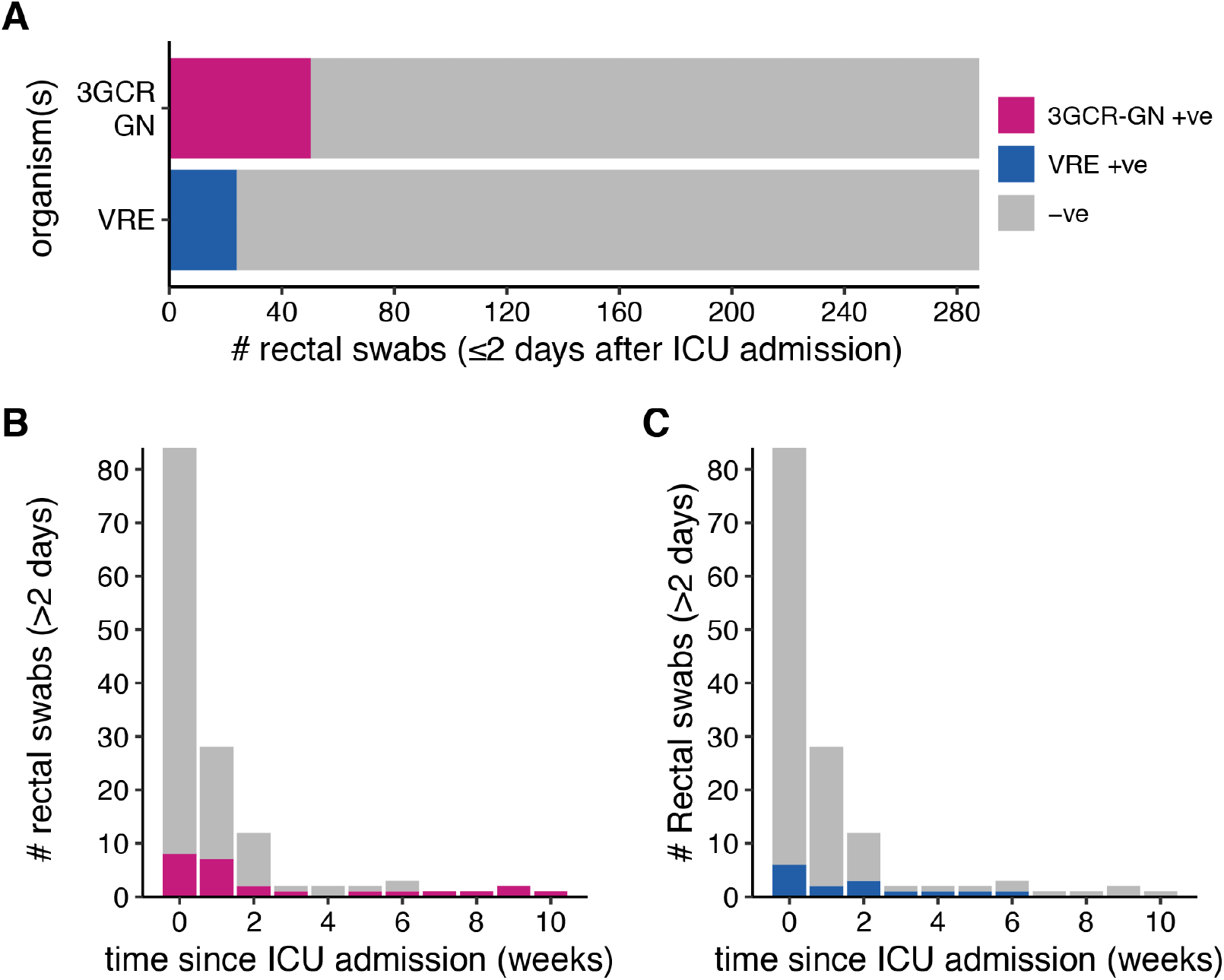
Prevalence of third generation cephalosporin-resistant Gram-negative (3GCR-GN) and VRE gut colonisation stratified by time since ICU admission. (**A**) Prevalence of 3GCR-GN organisms and VRE among patients swabbed within the first two days of ICU admission (baseline swabs). Prevalence of 3GCR-GN (**B**) and VRE (**C**) among patients, for swabs collected >2 days after ICU admission.

**Figure S3:**
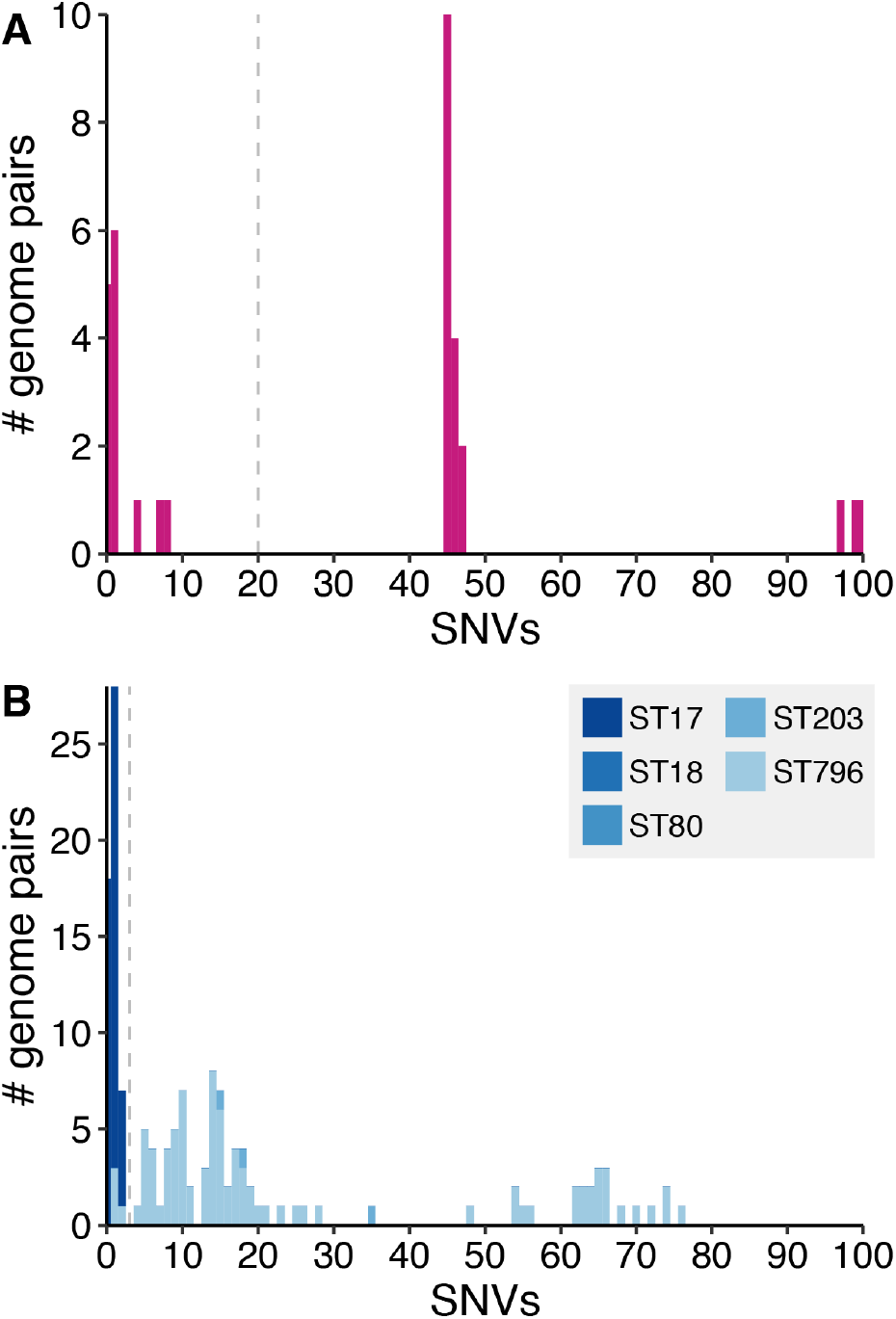
Distribution of single nucleotide variant (SNV) counts ≤100 for pairs of (A) third-generation cephalosporin-resistant Gram-negative (3GCR-GN) and (B) VRE isolates from different patients. Grey dashed lines indicate the SNV thresholds used to define putative transmissions (n ≤ 20 SNVs for 3GCR-GN and n ≤ 3 SNVs for VRE). Bars in (**B**) are coloured by the *E. faecium* multi-locus sequence type (ST) of the isolates as indicated in the legend.

